# INCIDENCE OF HEPATITIS B AND C VIRUSES AMONG THE SCAVENGERS IN KWARA STATE, NIGERIA

**DOI:** 10.1101/2022.01.26.22269849

**Authors:** Yusuf Olanrewaju Raufu, Adewoye Solomon Olayinka, Sawyerr Henry Olawale, Morufu Olalekan Raimi

**Affiliations:** Department of Environmental Health Science, Kwara State University, Malete, Nigeria; Department of Pure and Applied Biology, Ladoke Akintola University of Technology, Ogbomosho; Department of Community Medicine, Environmental Health Unit, Faculty of Clinical Sciences, Niger Delta University, Wilberforce Island, Bayelsa State, Nigeria

**Keywords:** Hepatitis B Virus, Hepatitis C Virus, Scavengers, Seroprevalence, Personal Protective Equipment, Kwara South, Kwara North, Kwara Central, Health Insurance Scheme

## Abstract

**Background:** Poor economic situations in developing nations had made scavenging a means of livelihood for millions of youth and women across the globe. Lack of proper source segregation of wastes in developing countries has increased the potential for the transmission of pathogens like Hepatitis B Virus (HBV) and Hepatitis C Virus (HCV).

**Objectives:** This study addressed issues relating to waste scavenging, the potential risk in waste scavenging work and seroprevalence of Hepatitis B and C Virus and its relationship with wastes scavenging among wastes scavengers in Kwara State.

**Methods:** A cross-sectional study was conducted among the scavengers working for major scrap dealers in the three senatorial districts of Kwara State. Consequently, to accomplish the objectives, both primary and secondary data sources were used. The primary data were collected via questionnaires, interviews, blood test and field observations. Whereas the secondary data were extracted from different published and unpublished materials. 240 respondents were administered with questionnaires and undergone hepatitis surface antigen test for both hepatitis B and C in nine of the sixteen local government area in the state. The data were analyzed using statistical package for social science (SPSS version 23) for descriptive and inferential at 5% level of significance. The prevalence of an HBV and HCV infection biological markers (HBsAg and HCsAg) and their associations with exposure to bio-medical waste, socio-demographic factors, and history of occupational injuries was examined.

**Results:** The seroprevalence of HBV and HCV infection among the scavengers were found to be 8.3% and 5% respectively indicating that scavengers are at risk of HBV and HCV infection.

**Conclusions:** It was discovered that there are incidences of Hepatitis B and C virus co-infection among scavengers. Also, awareness and compliance to the usage of PPE was found to be an important factor for protection scavengers against the virus. Therefore, vaccination against HBV, enforcement of usage of PPE, good hygiene practices, regular trainings on occupational safety, proper monitoring by regulatory agency and inclusion of scavengers in mandatory health insurance scheme will help to control risk of HBV and HCV infection among scavengers.

## Introduction

Wastes are inevitable remains from activities of human and animal. Daily activities of man entail constant discard of unwanted materials in form of wastes. Wastes management is therefore refers to the activities and actions required to manage waste from its inception to its final disposal [1]. While, municipal wastes are mixed and unsegregated at points of generation, undermining effective management when it comes to treatment and disposal practices. Indiscriminate disposal and sorting of mixed wastes by scavengers have the potential for increased environmental exposure to air pollution, toxic emissions from combusted or burnt municipal waste, proliferation of vermin and spread of diseases like hepatitis [2, 3, 4, 5, 6, 7, 8, 9, 10]. Hepatitis is an inflammatory condition of the liver and viral hepatitis is a conventional term used to denote hepatitis caused by hepatotrophic viruses Hepatitis A-G. High prevalence of these viruses especially hepatitis B is reported in Nigeria. Hepatitis B (alone accounting for nearly one million deaths annually) and C may cause liver cirrhosis and they can be contacted through contaminated blood and blood products [11].

Viral hepatitis has become a major health problem worldwide and cause acute and/or chronic hepatitis which can leads to the development of extensive liver scarring (cirrhosis), liver failure, liver cancer and death. As previous research has reported the endemic nature of viral hepatitis throughout sub-Saharan Africa and as a leading infectious killer globally, including its negative impact on human health [12, 13, 14, 15, 16, 17, 18, 19, 20, 21, 22, 23, 24]. While, viral hepatitis remains the tenth leading cause of death and the leading cause of liver cancer worldwide [25, 26]. It has also been suggested that hepatitis may increase the risk of pancreatic cancer [27]. Thus, the research is therefore, aimed at finding out the seroprevalence of hepatitis B, hepatitis C and co-infection with Hepatitis B and C among scavengers in Kwara State, Nigeria. It is anticipated that the findings from this study will necessitate the need for thorough screening of blood among scavengers in Kwara State toward reducing risk among scavengers in Nigeria. The scope of the work is however narrowed to nine local government areas out of sixteen local government areas in the state. The respondents are therefore chosen in three local governments from each senatorial district of Kwara North, Kwara South and Kwara Central respectively.

## Methodologies

### Study Area

Kwara state is located in the North Central Nigeria; it lies between 11°2 and 11°45 North and Longitude 2°45 and 64 East (see figure 1 below) [4, 8, 10]. The state covers a land area of 35,705 square kilometers and has a population of 2,371,089 [27]. It has 16 Local Government Areas and the population of 2.37 million people according to 2006 census.

**Figure 1:**
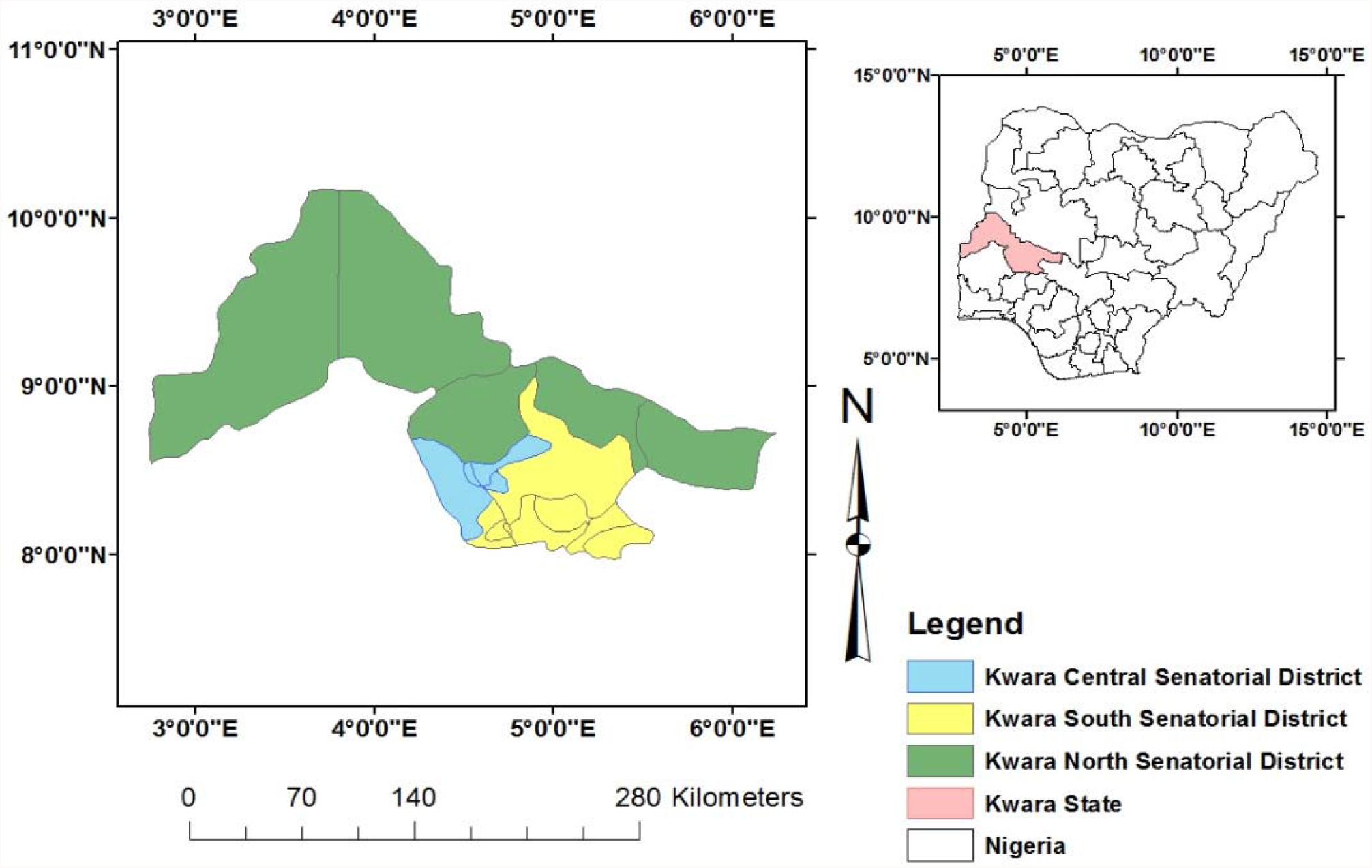
Map of Nigeria Showing Kwara State and the study areas.

### Study Population

The study population were scavengers working with major scrap dealers in Kwara State as well as on the dumpsites across the state.

### Sampling Techniques

Scavengers were selected from the registered scrap dealers through the State Environmental Protection Agency (KWASEPA) as well as the association of scrap dealers working on the dumpsites. Respondents were administered with questionnaires and their consents were sought before proceeding to the second stage (Hepatitis B and C Screening) of the study. The appropriate sample size was determined using fishers sample size formula, the total sample size was determined as 240 including 10% non-response rate.

### Sample Collection

Blood samples were collected with the help of a medical laboratory scientist from respondents at three dumpsites in Ilorin Metropolis using intravenous needles. The needles and syringes used for the collection of blood samples were dried and sterile. The serum obtained was tested for hepatitis B surface antigen (HBsAg) and anti-hepatitis C antibodies using a Diaspot rapid diagnostic test strip.

### Rapid Diagnostics Test

The Diaspot rapid diagnostic test is used to qualitatively detect the presence of HBsAg and HCsAg in serum or plasma specimens. The test utilizes a combination of monoclonal and polyclonal antibodies to selectively detect elevated levels of HBsAg and HCsAg in serum or plasma. During testing, the serum or plasma specimen reacts with the particles coated with anti-HBsAg and anti-H anti-HCV antibody. The mixture migrates upward on the membrane chromatographically by capillary action to react with anti-HBsAg and anti-HVC antibodies on the membrane and generate a colored line. The presence of the colored line in the test region indicates a positive result, while its absence indicates a negative result. To serve as a procedural control, a colored line will always appear in the control line region indicating that the proper volume of specimen has been added and membrane wicking has occurred. The manufacturers’ instructions were strictly followed in the performance of these tests. The test strips, serum or plasma specimens were allowed to equilibrate to room temperature (15-30° C) prior to testing. The test device was placed on a clean, level surface and 60 μl of serum or plasma was added to the sample well of the device. The sample rehydrated and was mixed with the red colloidal gold conjugate, which flowed into the membrane.

### Data Instrument

A semi-structured, interviewer administered questionnaire was used to elicit information on the socio-demographics and occupational hazards of all respondents. The instrument was pre-tested using 24 scavengers at Amoyo town from a group similar to the main study group. Each question was translated into the local languages (i.e. Yoruba, Nupe, Fulani and Hausa) for those that could not read English, to help the respondents to give true and accurate answers.

### Data Analysis

Data was analyzed using SPSS 23, using descriptive statistic such as mean, frequencies, charts and graphs. Also, inferential statistic using six different statistical tests were used for the analysis. The tests used are Pearson chi-square, likelihood ratio, linear-by-linear association, phi, Cramer’s, and contingency coefficient using the level of significance (0.05).

### Ethical Approval

Ethical clearance for this study was obtained from the Kwara State Ministry of Health Ethical Review Committee. Informed consent of participants were sought before taking part in the study.

## Results

### Socio-Demographic Characteristics of scavengers

Here we present the results of the wastes scavengers based on their age, marital status, educational level and tribal status respectively.

### Source: Author field survey, 2021

The Table 1 above shows results from wastes scavengers’ socio-demographic data. While, figure 1 below indicate the incidence of hepatitis B virus among scavengers across the three senatorial districts in Kwara State. The results of hepatitis surface antigen test showed twenty-three (23) scavengers which equivalent 9.6% tested positive for hepatitis B across the three senatorial districts. The Kwara central senatorial district has the highest hepatitis B positive rate with ten (10) hepatitis B positive cases. Kwara south senatorial districts followed with seven (7) hepatitis B positive incidences and Kwara north senatorial districts has the least cases of hepatitis B positivity with six (6) cases. Figure 2 shows the Hepatitis C surface antigen screening for the scavengers and the results of the screening is as presented in the chart below. The overall percentage of hepatitis C positivity in the state was 5% and the results also show that Kwara central has the highest incidences of hepatitis C with seven (7) cases of positivity. Kwara south hepatitis C positivity is next to Kwara central with three (3) cases and Kwara north maintained the least incidence of hepatitis C positive cases with just two (2) incidences. In the Figure 3, co-infection of scavengers with both hepatitis B and hepatitis C were examined and it was revealed that there are cases of scavengers being infected with both hepatitis B and hepatitis C. The chart below present the results of incidences of co-infection by scavengers.The results revealed there are seven (7) co-infection among the scavengers which constitutes 2.91% of the total scavenger population. It was revealed that Kwara central has the highest number of co-infection of hepatitis B and C with four (4) positive cases among the respondents while the two senatorial districts of Kwara south and Kwara north has two (2) cases of co-infection each. Figure 4 shows compliants of wastes scavengers to the use of personal protective equipment (PPE). Compliance to the use of PPE was determined by the frequency of the PPE usage. Those that make use of PPE for at least five days in a week are categorised to have complied with the use. Kwara south senatorial districts lead in terms of the usage of PPE with 48.5% compliants while Kwara central closely followed with 43.1% of their scavengers in the district. The least compliants was found in Kwara north with 33.8% of scavengers in Kwara north complied with PPE usage.

**Table 1:** Socio-demographic Characteristics of Respondents.

| **AGE** | **Frequency** | **Percent** |
| --- | --- | --- |
| 10 – 20 | 61 | 25.4 |
| 21 – 30 | 109 | 45.4 |
| 31 – 40 | 67 | 27.9 |
| 41- above | 3 | 1.3 |
| Total | 240 | 100.0 |
| **GENDER** | **Frequency** | **Percent** |
| Male | 231 | 96.2 |
| Female | 9 | 3.8 |
| Total | 240 | 100 |
| **TRIBE** | **Frequency** | **Percent** |
| Hausa | 194 | 80.8 |
| Nupe | 8 | 3.3 |
| Yoruba | 26 | 10.8 |
| Igbo | 3 | 1.3 |
| Fulani | 8 | 3.3 |
| Others | 1 | 0.4 |
| Total | 240 | 100.0 |
| **MARITAL STATUS** | **Frequency** | **Percent** |
| Single | 129 | 53.8 |
| Married | 111 | 46.2 |
| Total | 240 | 100 |
| **EDUCATIONAL QUALIFICATION** | **Frequency** | **Percent** |
| None | 68 | 28.3 |
| Primary level | 93 | 38.8 |
| Secondary level | 42 | 17.5 |
| Tertiary level | 5 | 2.1 |
| Islamiyah | 32 | 13.3 |
| Total | 240 | 100.0 |

**Source: Author field survey, 2021**

**Figure 2:**
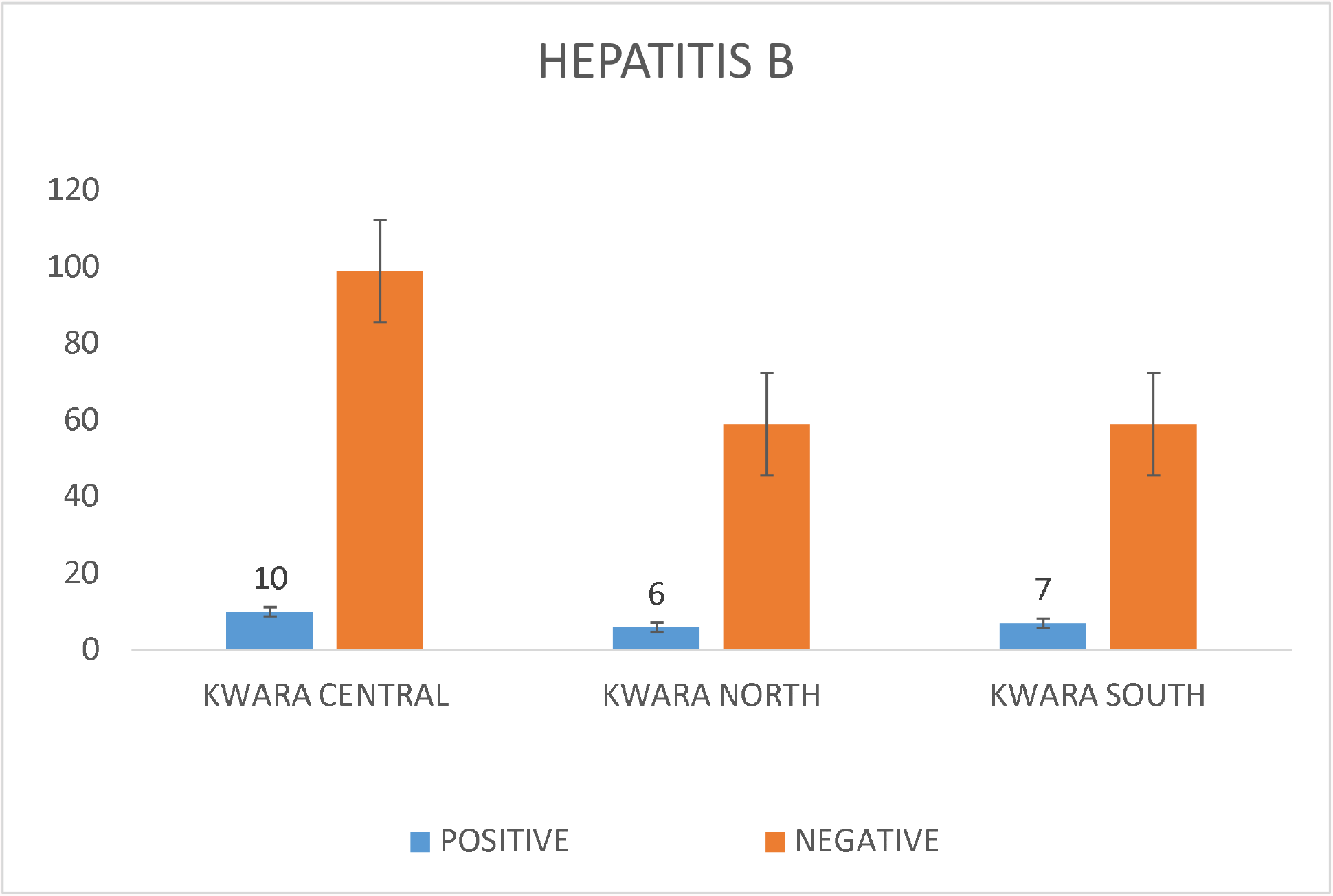
Hepatitis B incidences among scavenger across the three senatorial districts.

**Figure 3:**
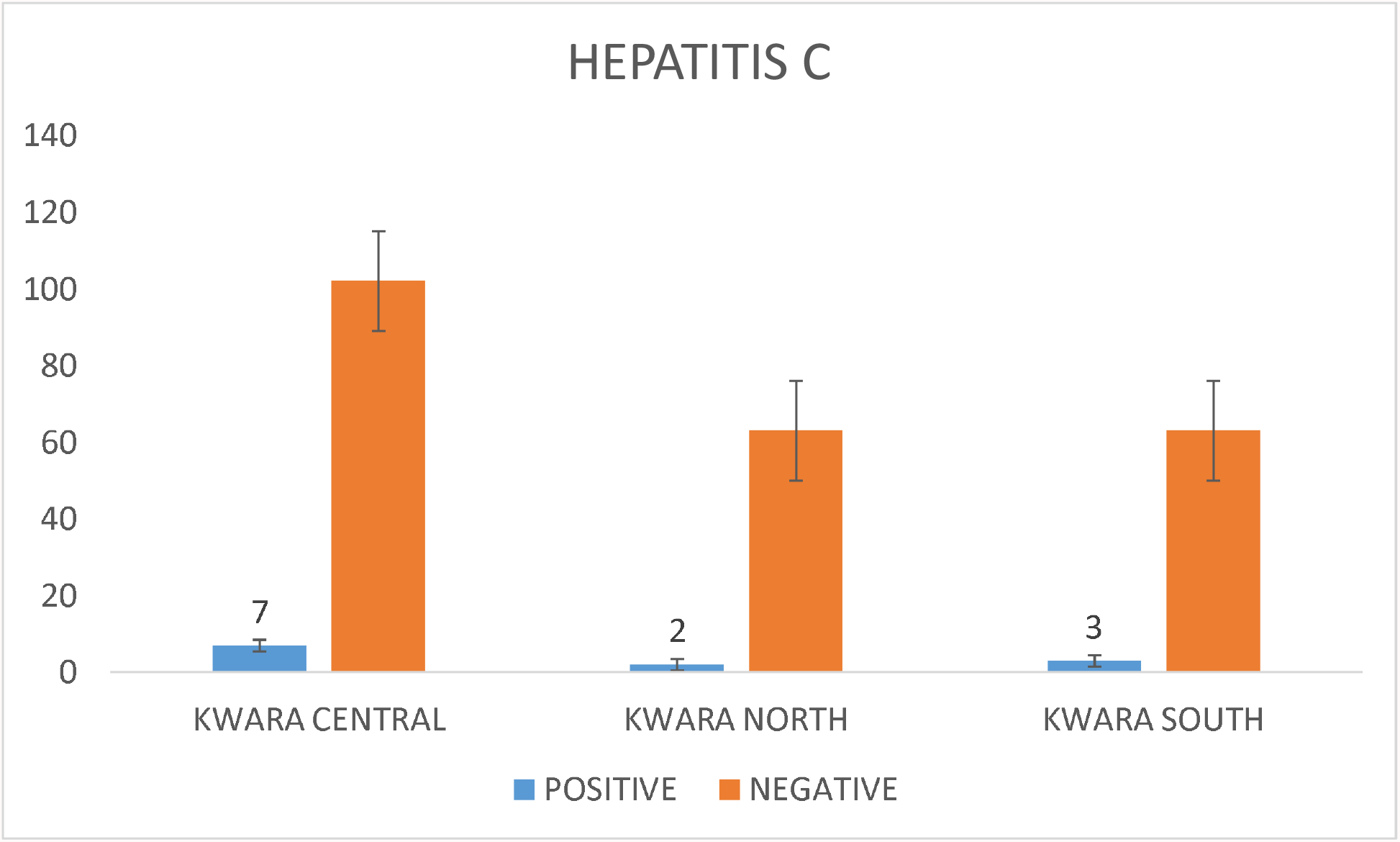
Hepatitis C incidences among scavenger across the three senatorial districts.

**Figure 4:**
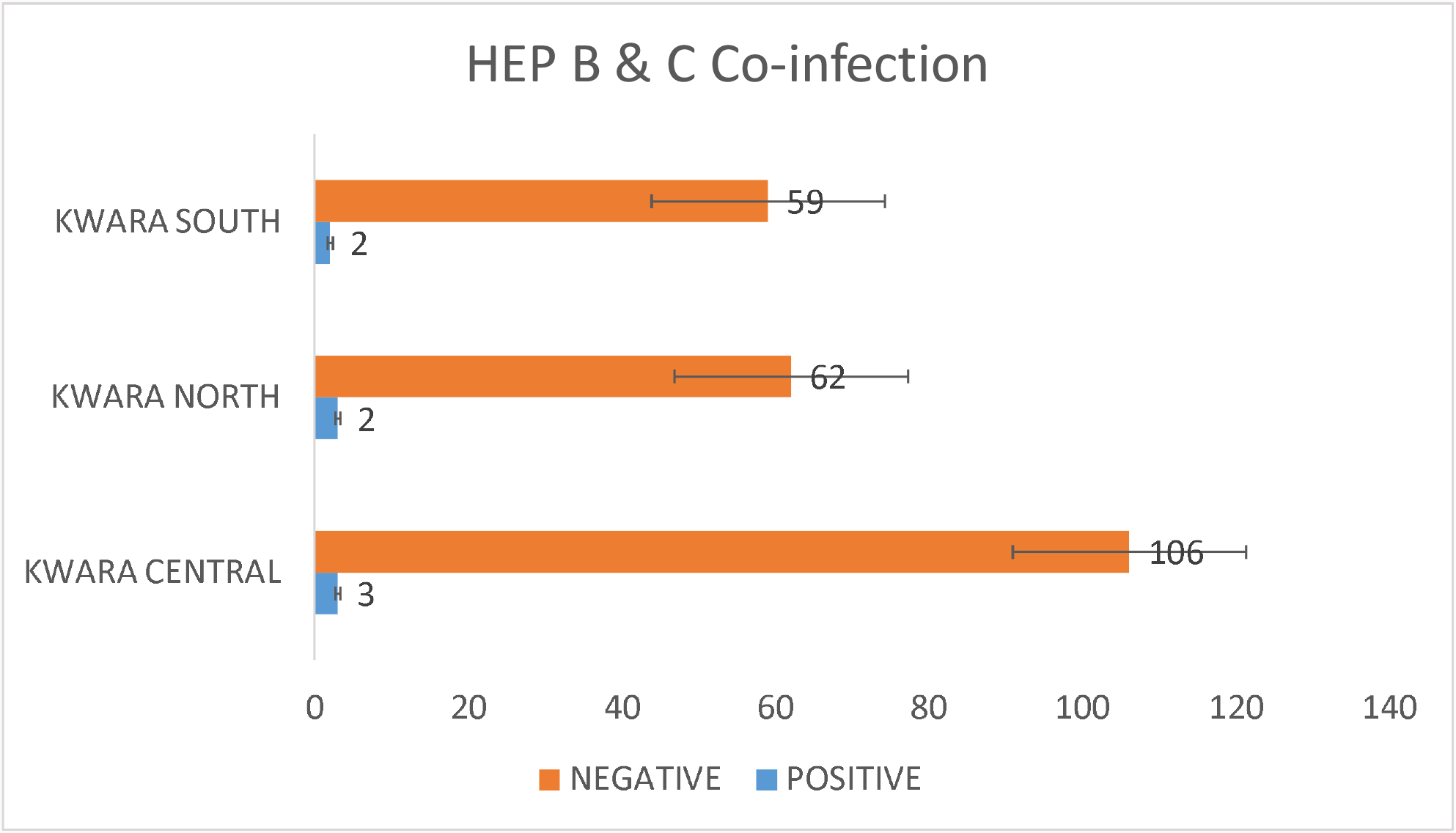
Co-infection of Hepatitis B and C incidences among scavenger across the three senatorial districts.

**Figure 4:**
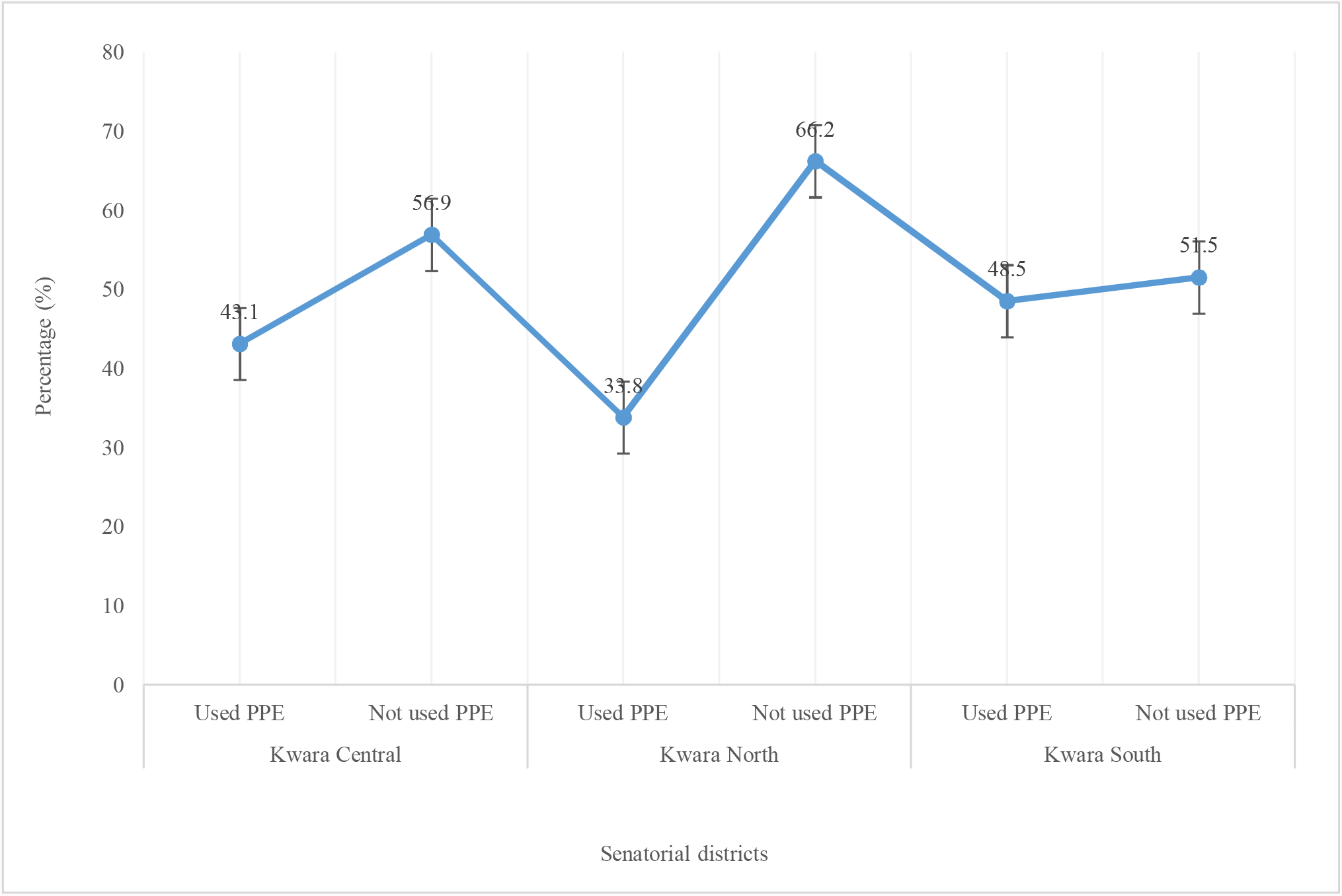
Compliant to PPE usage across the three senatorial districts.

## Discussion

Nigeria is among the countries with a high burden of viral hepatitis with a Hepatitis B Virus (HBV) and Hepatitis C Virus (HCV) prevalence of 11% and 2.2%, respectively [28]. This study revealed that majority of the scavengers in the study are in the age bracket 21-30 years which is 45.4% that is in agreement with Irabor *et al* [29] which concluded that the highest population of scavengers i.e., 36% are in the age bracket 20 - 29 years and Isaac *et al* [30] stated that majority 35.42% of respondents are between 18-28 years. In terms of education, majority of wastes scavengers in Kwara state all to the category of primary school leavers as the constitute 38.8% of total respondents and closely followed by those without any formal education which constitute 28.3%. Secondary school certificate holders constitute 17.5% while only 2.1% of scavengers has a higher educational certificate. This is in agreement with Adebola *et al* [31] in his work on wastes worker and scavengers where he found that majority (51.1%) of scavengers in Ibadan are primary school leavers. Scavenging job was found to be a male dominated occupation because 93.6% of respondents in this study are male scavengers which agrees with the work of Oteng-Ababio [32] which stated that about 90% of the scavenger population are male while female scavengers constitute approximately 10% in Kumasi. Similarly, in another study male made up 86% of e-waste scavengers found at the Agbogbloshie e-waste scrap yard in Accra as reported by Rankokwane and Gwebu, [33] and Afon [34]. It was also discovered that majority of scavengers are immigrant from the northern part of the country and this is because 80.8% of respondents of this study are Hausa by tribe and this agrees with Pinto [35] that asserted that majority of scavengers are immigrants. In terms of marital status, it was found that 53.8% of the scavengers are still unmarried and this three (3) can be as a result of high population of young adults that constitute majority of respondents. The research findings show incidence of hepatitis B and hepatitis C and co-infection with hepatitis B and C among wastes scavengers in Kwara state. Seroprevalence of hepatitis B detected through the use of hepatitis B test strip. Micro point brand was used for rapid test of the blood samples of scavengers. The results of hepatitis surface antigen test showed twenty-three (23) scavengers which equivalent 9.6% tested positive for hepatitis B. Across the three senatorial districts, Kwara central has the highest hepatitis B positive rate with ten (10) hepatitis B positive cases. Kwara south followed with seven (7) hepatitis B positive incidences and Kwara north has the least cases of hepatitis B positivity with six (6) cases. This is in agreement with the work of Yusuf *et al* [36] which also shows a high hepatitis B positivity among scavengers in Ilorin metropolis. Also, the result corroborates Sawyerr *et al* [2] in their findings that revealed higher incidence of hepatitis B virus among wastes scavengers in comparison with the municipal street sweepers. Hepatitis C screening for the scavengers and the results show the overall percentage of hepatitis C positivity in the state to be 5% and the results also indicated that Kwara central has the highest incidences of hepatitis C with seven (7) cases of positivity. This can be as a result of higher population of the state capital and proliferation of illegal dumpsites. Kwara south hepatitis C positivity is next to Kwara central with three (3) cases and Kwara north maintained the least incidence of hepatitis C positive cases with just two (2) incidences. This incidence of hepatitis C virus is higher than the national average occurrences in non-wastes workers as reported to be 2.2%, [28]. Also, in another work by Agbede *et al*. [37] which stated that the HCV prevalence of 1.4% was seen among mothers in Ilorin metropolis. Though this research supported the findings of Adewuyi [38] who reported 5% prevalence of antibodies to hepatitis C virus among normal blood donors and multi-transfused sickle-cell anaemia patients in the same environment. The distribution of co-infection of scavengers with both hepatitis B and hepatitis C across the state was 7 (2.92%) co-infection. This result is very close to the findings of Taiwo *et al*. [39] in their study on patients in Lagos State University Teaching Hospital (LASUTH), where it was reported that Dual presence of HBsAg and anti-HCV was observed in 4(3.9%). It was revealed that Kwara central came top with 3 (42.8% of positive) co-infection cases while Kwara south and Kwara north has 2 (28.6% of positive) co-infection cases.

## Conclusion

This study revealed that the seroprevalence of hepatitis B among scavengers in the state which 9.6% is still under the national average but hepatitis C was found to be 5% which is greater than the national average. It is therefore recommended that enabling law should be promulgated to regulate scavenging and make mandatory data capture of all scavengers. This will help the Government to know the population of scavengers and make adequate provision for their enlightenment and intervention that reduce the health burden of scavengers. Government agencies like the Kwara Environmental Protection Agency (KWASEPA) and the Ministry of Environment and Forestry should ensure that scavengers are adequately making use of PPE with strict enforcement measures. Free vaccination against HBV should be provided for scavenger, educational campaigns and regular training on occupational health and safety programs and health surveillance should be instituted for all waste scavengers with emphasizes on good work practices and personal hygiene practices.

### Study Limitations

The small number of samples used in this study could place a limit to the outcome of this study. This is largely attributed to limited funds as there is no grant and sponsorship from any agencies.

## Data Availability

All data produced in the present work are contained in the manuscript

## Acknowledgements

The authors remain sincerely grateful to all scavengers that participated in the study and also all medical laboratory scientists and research assistant who provided logistic support during this study.

## Data availability

The data used to support the findings of this study are included in the article. The raw data of this study will be made available on request. All requests should be made to the corresponding author of this article.

## Scavenger consent

*Obtained*

## Ethics approval

*Ethical approval was granted from the Kwara State Ministry of Health Ethical Review Committee. Informed consent was obtained from each respondent*.

